# Validation of a Composite Mortality Endpoint in a Large, Clinico-Genomic Real-World Database of Patients with Advanced Cancer

**DOI:** 10.1101/2025.08.20.25334011

**Authors:** Joshuah Kapilivsky, Farahnaz Islam, Emma K. Roth, Jessica Dow, Shannon Moran, Emilie Scherrer, Seung Won Hyun, Chithra Sangli

## Abstract

**Purpose:** Real-world data (RWD) from electronic health records (EHRs) and next-generation sequencing are increasingly used to study treatment effectiveness in molecularly refined patient populations. Incomplete mortality data in EHR can overestimate survival rates in RWD studies. While the National Death Index (NDI) is the gold standard for mortality data in the United States, its limited accessibility and reporting delays hinder timely research. Instead, EHR datasets are often supplemented with external mortality data sources to improve mortality data capture. This study evaluated a composite mortality variable against NDI records using a large cohort of advanced cancer patients from a real-world oncology database.

**Methods:** De-identified clinical and molecular data from patients with advanced solid tumors were linked with third-party mortality and claims datasets using deterministic tokenization. Vital status and death dates were harmonized across sources. Patient identifiers were submitted to NDI, and true matches were de-identified and joined for analysis. Performance metrics (sensitivity, specificity, positive predictive value [PPV], negative predictive value [NPV]) were calculated using NDI as ground truth. Date agreement was assessed at 0, ±15, and ±30-day tolerances. Subgroup analyses and a cumulative cases/dynamic controls (CC/DC) approach were also performed.

**Results:** Among 17,597 patients, the composite mortality variable demonstrated 82% sensitivity and 95% specificity against NDI. PPV was 96%, and NPV was 77%. Exact date agreement was 86%, increasing to 94% within a ±15-day tolerance and 96% within a ±30-day tolerance. Incorporating third-party mortality and claims data substantially improved sensitivity from 17% (EHR alone) to 82%. Sensitivity remained stable across subgroups but showed variation by age, cancer type, geographic region, and race. With the CC/DC approach, sensitivity was 96% at 6 months, 97% at 12 months, and 98% at 24 months, with specificity above 98% across these timeframes.

**Conclusions:** The composite mortality variable is a robust, reliable endpoint for real-world evidence analyses. Its high accuracy for identified deaths and appropriate censoring of lost-to-follow-up patients support its use in overall survival analyses. This validation is a foundational step towards high-quality research to improve patient outcomes and advance cancer drug development using this multimodal dataset.

**Clinical trial number:** not applicable

## Background

Real-world data (RWD), derived from sources like electronic health records from routine clinical care, provide a vital complement to evidence generated from traditional clinical trials. These data allow researchers to study treatment effectiveness in real-world populations that may have more diverse clinical characteristics than what can practically be studied in a clinical trial. With the development and adoption of modern targeted therapies in oncology, it is also increasingly important to integrate molecular data from next-generation sequencing (NGS) into these clinical datasets to investigate relationships between specific genomic alterations and patient outcomes. A common method for assessing treatment effectiveness in this context is overall survival (OS) analysis, which fundamentally depends on accurate mortality data. Therefore, the full potential of these rich clinico-genomic datasets can only be realized if their mortality data are accurate and reliable.

In the United States, the National Death Index (NDI) is widely regarded as the gold standard for mortality data due to its comprehensive coverage. However, the significant time lags, logistical hurdles, and use limitations involved in accessing NDI data can create a bottleneck, limiting its utility for the pace and volume of research required to advance precision oncology. Alternative data sources, such as electronic health records (EHRs), are more readily accessible but are often incomplete, as patients may transfer their care to other institutions and become lost to follow-up (1,2). Relying solely on EHR data can therefore lead to under-ascertainment of death events and an overestimation of survival rates, skewing research findings (3,4).

To address these limitations, a common strategy is to create a composite mortality variable by integrating data from multiple sources. Numerous studies have shown that combining EHR data with information from third-party mortality databases, which aggregate data from sources like the Social Security Death Index, public records, and obituaries, can significantly improve the sensitivity and completeness of mortality capture (4–7). Therefore, while such a composite approach may not achieve the same level of completeness as the NDI, the advantages of reduced data latency and improved accessibility can provide a mortality endpoint better suited to the pace and volume of research needed to advance precision oncology.

In this study, we evaluated the performance of the composite mortality variable used within the Tempus real-world multimodal database. To validate this endpoint, we benchmarked its performance against the NDI in a large cohort of patients with advanced cancer who underwent Tempus sequencing as part of their routine clinical care.

## Methods

### Study Design

This study was a non-interventional retrospective analysis of mortality data from the Tempus real-world multimodal database. The objective was to validate the Tempus composite mortality variable by benchmarking it against death information from the NDI.

### Data Sources

The Tempus multimodal database consists of de-identified longitudinal clinical and molecular data for cancer patients who undergo sequencing at Tempus. Tempus offers oncology sequencing services to roughly 65% of academic medical centers and several hundred community institutions in the U.S. Clinical histories provided in furtherance of testing are principally obtained from EHRs or other electronic systems. After being de-identified, records are supplemented with third-party de-identified datasets, including an administrative claims dataset and a mortality dataset provided by Veritas Data Research. The Veritas Fact of Death Mortality Data Index is a composite dataset of U.S. mortality records from more than 40,000 sources, including the Social Security Administration’s Limited Access Death Master File (SSA LADMF), obituary notices, public cemetery records, state death registries, and medical claims.

External de-identified datasets are joined to Tempus’s de-identified data at the patient grain using privacy-preserving, deterministic patient linkages. Each organization (i.e., Tempus and the third-party data provider) uses an irreversible, cryptographic process to generate a unique, non-identifying code, known as a “token”, from Personally Identifiable Information (PII) available in their records. Use of such a process prevents use of the token alone to reconstruct the original PII. The process is also deterministic: the same set of PII will always follow the same cryptographic steps. Each organization uses its own secret key during token generation, which ensures the resulting tokens are unique to that organization. This prevents tokens from being matched between organizations without an explicit agreement. Tempus and its third party data providers use the same tokenization vendor, and when Tempus agrees with a data vendor to exchange data, the tokenization vendor provides each party with the necessary encryption and decryption keys to convert their organization-specific tokens into a shared token format unique to that partnership. The data vendor can then send data to Tempus using the shared token format, and Tempus can convert to its own tokens to identify de-identified records of common patients and create integrated, de-identified patient records (8).

The National Death Index (NDI) is a centralized database maintained by the National Center for Health Statistics (NCHS) that provides death record information sourced from death certificates. According to the Centers for Disease Control (CDC), it is the most complete source of death information in the United States (9).

For this study, patient identifiers—including name, date of birth, sex, and, when available, Social Security Number—were submitted to the NDI via secure file transfer. The NDI returned potential matches, and true matches were selected based on the recommendations in the NDI User’s Guide (10). Only matches meeting these criteria were considered true deaths; subjects without a true death recorded in NDI were considered alive. After matching, NDI results were de-identified and joined to the corresponding Tempus record for the limited purposes of performing analysis in support of this study.

### Cohort Selection (Figure 1)

Patients were eligible for inclusion in this study if they had a primary diagnosis of non-small cell lung cancer (NSCLC), breast cancer (BC), ovarian cancer (OC), pancreatic cancer (PANC), colorectal cancer (CRC), or prostate cancer (PC) between 2016 and 2020. These specific cancer types were chosen because they represent the predominant solid tumor populations within the Tempus multimodal database. A diagnosis period of 2016-2020 was selected to guarantee at least two years of mortality ascertainment through the National Death Index (NDI). At the time of NDI data submission, NDI records were available through December 31, 2022.

Patients must have been 18 or older at the time of cancer diagnosis. To enrich for death events, only patients with locally advanced, unresectable, or metastatic disease stages were included, as defined by disease-specific staging criteria. Additional inclusion criteria required that patients have sufficient personally identifiable information (PII) available for linking with third-party datasets and for submission to the NDI.

Initial patient eligibility was determined prior to submission to NDI. After receipt from NDI, patient eligibility was re-confirmed with refreshed data from the live database and subjects rejected from the NDI search were excluded. The study dataset was then frozen and all analyses were performed using the frozen dataset.

### Vital Status Harmonization

All data sources in the integrated patient record were combined into a single high-confidence, internally-consistent representation of vital status (i.e., dead/alive). First, all dates that a patient was known to be alive were extracted using a predefined list of fields in the data model that have a strong semantic relationship with alive status. For example, the administration of an infusion or an inpatient hospital admission indicate that a patient is alive, but an automated prescription fill in EHR or the date a claim was submitted to a payer may actually take place after a patient’s date of death and thus do not indicate aliveness. The latest date from these alive dates was selected as each patients’ last known alive date. Next, each death date available in the integrated clinical record was extracted and compared against the last known alive date. Any death date that precedes the last known alive date was discarded. Death dates that remain were ranked by the reliability of their source to select up to one mortality date per patient.

At the time of this study, the routine NDI data file included deaths through the end of 2022. To enable fair comparison against NDI, Tempus vital status data were truncated to match. Patients with a Tempus last known alive date or deceased date after December 31, 2022 were considered alive and their last known alive date was changed to December 31, 2022.

### Validation of the Tempus Composite Mortality Variable

For each patient, vital status (alive or deceased) and date of death were compared between the Tempus dataset and the NDI, with NDI considered as ground truth. Sensitivity was defined as the proportion of NDI-classified deaths that were also identified as deceased in the Tempus dataset. Specificity was defined as the proportion of NDI-classified living patients who were not identified as deceased in the Tempus dataset. PPV and NPV were calculated as the proportions of Tempus-classified deaths and living patients, respectively, that were confirmed by the NDI. Exact 95% confidence intervals for all performance metrics were calculated using the Clopper-Pearson method.

Date agreement was assessed by calculating the proportion of Tempus death dates that matched NDI deaths with identical dates of death, as well as within ±15-day and ±30-day windows. Death dates in the Tempus database that fall within the allowed time window from the NDI death date were counted as an agreement. Death dates that do not fall within the time window were counted as a disagreement. Patients with a Tempus death date without an NDI death date were also counted as a disagreement.

While the majority of Tempus death dates are available from the source at year-month-day resolution, some death dates are available at only year-month resolution. These year-month dates are imputed to year-month-day resolution as part of Tempus’s standard de-identification process. All NDI death dates are available at year-month-day resolution. Only Tempus death dates available at year-month-day resolution were included in exact day and ±15-day agreement metrics. ±30-day metrics included Tempus death dates at either year-month-day or year-month resolution.

The contribution of individual mortality data streams was assessed by calculating the metrics described above on individual mortality data streams (i.e., EHR, third-party mortality, third-party claims), as well as upon addition of each data stream (i.e., EHR, EHR + third-party mortality, EHR + third-party mortality + third-party claims). Exploratory subgroup analyses were conducted by age at diagnosis, cancer type, geographic region, year of diagnosis, number of recorded lines of therapy, race, and sex. Additionally, sensitivity and date agreement metrics were reported stratified by year of death according to NDI. Because this stratification focuses solely on patients with NDI-classified deaths, components necessary for calculating specificity, PPV, and NPV (i.e., true negatives and false positives) are eliminated. Therefore, these metrics were not reported for this stratification variable.

The endpoints above assess the completeness of Tempus’s capture of all deaths in the NDI, treating any missed death event as a false negative. However, in real-world overall survival analyses—such as those using Kaplan-Meier estimation or Cox proportional hazards models— Tempus mortality data are typically interpreted alongside the last known alive date, with patients classified as deceased, alive, or censored. To provide additional context on how Tempus data reflects patients without a recorded date of death, we conducted a naive estimation of sensitivity and specificity at t = 6, 12, and 24 months using the cumulative cases / dynamic controls (CC/DC) approach (11). This method restricts the analysis to patients with sufficient follow-up to be evaluable for survival at each time point by excluding those censored prior to time t.

### Data Availability

Data used in this research were collected in a real-world healthcare setting and are subject to controlled access for privacy and proprietary reasons. Furthermore, access to and use of National Death Index (NDI) data in this study are governed by a formal agreement signed as part of the NDI application process. This agreement stipulates that the data may not be published or released in any other form and are to be utilized exclusively for the objectives of this specific study (10).

### Ethics Statement

All data collected and used complied with the requirements and standards of the Health Insurance Portability and Accountability Act (HIPAA). Institutional Review Board (IRB) approval of the study protocol was obtained. This was a noninterventional study using routinely collected data, and informed consent was waived by the IRB (Advarra Pro00076789). This study was also reviewed and approved by staff from the NDI.

## Results

### Study Population

A total of 17,597 patients with advanced or metastatic solid tumors (NSCLC, BC, OC, PANC, CRC, and PC) diagnosed between 2016 and 2020 were included in the evaluable cohort (**Figure 1**). All patients were aged ≥18 years at diagnosis and had sufficient identifiers for National Death Index (NDI) matching and third-party data integration.

**Fig 1.**
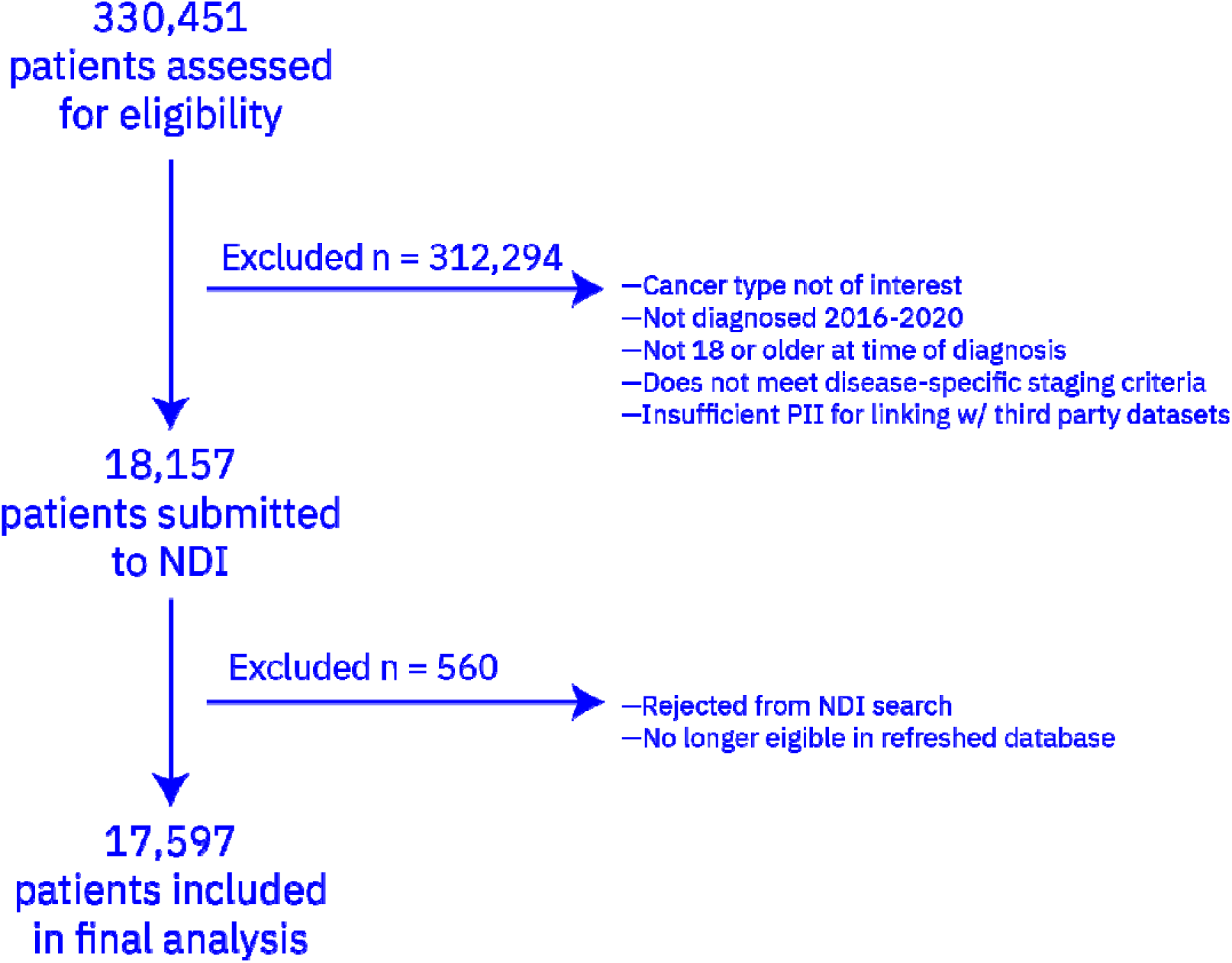
Cohort funnel

### Primary Endpoint (Table 1)

When benchmarked against the NDI as the gold standard, the Tempus composite mortality dataset demonstrated a sensitivity of 82%, indicating that 82% of deaths recorded in the NDI were also captured in the Tempus dataset. The specificity was 95%, reflecting a low rate of false positive death assigned to patients not classified as deceased in the NDI. The positive predictive value was 96% and the negative predictive value was 77%.

**Table 1.** Primary Endpoint Results

| Number of Subjects | Sensitivity | Specificity | Positive Predictive Value | Negative Predictive Value | Date Agreement (0 Day Tolerance) | Date Agreement (15 Day Tolerance) | Date Agreement (30 Day Tolerance) |
| --- | --- | --- | --- | --- | --- | --- | --- |
| 17597 | 81.79%<br>(81.05%, 82.52%) | 94.82%<br>(94.26%, 95.33%) | 96.19%<br>(95.78%, 96.57%) | 76.51%<br>(75.59%, 77.42%) | 86.11%<br>(85.37%, 86.82%) | 93.78%<br>(93.26%, 94.27%) | 95.61%<br>(95.17%, 96.02%) |

### Date Agreement

Death dates available as full precision (YYYY-MM-DD) in Tempus data had an exact date match with NDI of 86%. Date agreement increased to 94% when allowing for a ±15 day tolerance. For dates available as full precision (YYYY-MM-DD) or year-month precision (YYYY-MM), 96% matched to NDI within a ±30 day tolerance.

### Combining Mortality Datasets (Table 2)

Analysis of individual mortality data streams revealed that the composite approach substantially improved death capture. Without the inclusion of third-party datasets, only 17% of NDI deaths would have been captured using Tempus EHR-derived data alone. The addition of third-party mortality data increased sensitivity to 80%, and further addition of third-party claims data increased sensitivity to 82%. Specificity was 99% for Tempus EHR-derived data alone, and decreased to 95% with the addition of mortality sources. The positive predictive value was relatively stable with the addition of mortality sources, starting at 97% for Tempus EHR-derived data alone and decreasing to 96% for the Tempus Composite Mortality Variable. Negative predictive value for Tempus EHR-derived data alone was 43% and increased to 77% with the inclusion of additional sources. For exact date agreement, we observed an increase from 80% with Tempus EHR-derived data alone to 86% with the composite variable. The ±15 day agreement similarly increased from 93% to 94%, and ±30 day agreement remained stable at 96%.

**Table 2:**
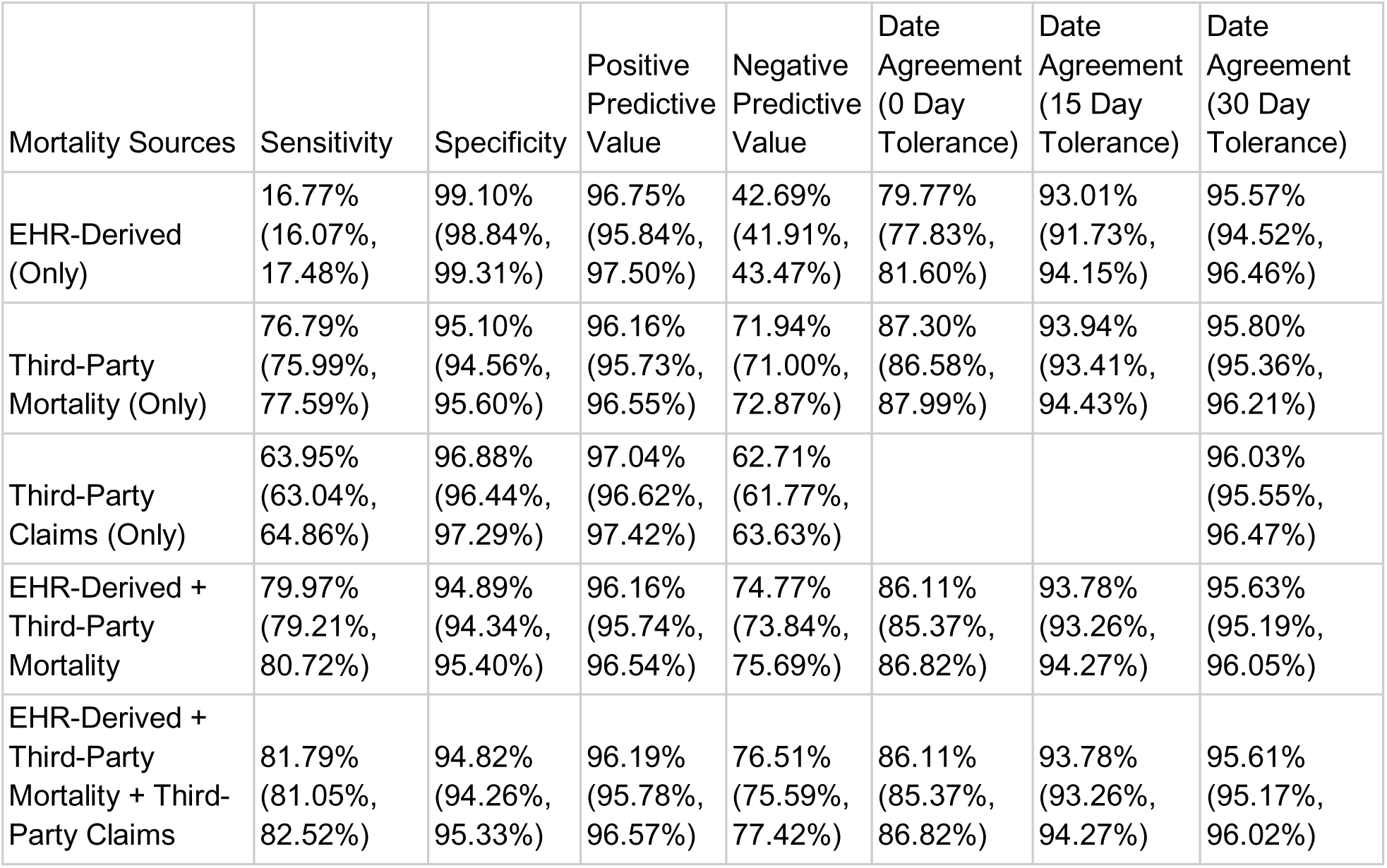
Performance Metrics by Mortality Data Stream(s)

| Mortality Sources | Sensitivity | Specificity | Positive Predictive Value | Negative Predictive Value | Date Agreement (0 Day Tolerance) | Date Agreement (15 Day Tolerance) | Date Agreement (30 Day Tolerance) |
| --- | --- | --- | --- | --- | --- | --- | --- |
| EHR-Derived (Only) | 16.77%<br>(16.07%, 17.48%) | 99.10%<br>(98.84%, 99.31%) | 96.75%<br>(95.84%, 97.50%) | 42.69%<br>(41.91%, 43.47%) | 79.77%<br>(77.83%, 81.60%) | 93.01%<br>(91.73%, 94.15%) | 95.57%<br>(94.52%, 96.46%) |
| Third-Party Mortality (Only) | 76.79%<br>(75.99%, 77.59%) | 95.10%<br>(94.56%, 95.60%) | 96.16%<br>(95.73%, 96.55%) | 71.94%<br>(71.00%, 72.87%) | 87.30%<br>(86.58%, 87.99%) | 93.94%<br>(93.41%, 94.43%) | 95.80%<br>(95.36%, 96.21%) |
| Third-Party Claims (Only) | 63.95%<br>(63.04%, 64.86%) | 96.88%<br>(96.44%, 97.29%) | 97.04%<br>(96.62%, 97.42%) | 62.71%<br>(61.77%, 63.63%) |  |  | 96.03%<br>(95.55%, 96.47%) |
| EHR-Derived + Third-Party Mortality | 79.97%<br>(79.21%, 80.72%) | 94.89%<br>(94.34%, 95.40%) | 96.16%<br>(95.74%, 96.54%) | 74.77%<br>(73.84%, 75.69%) | 86.11%<br>(85.37%, 86.82%) | 93.78%<br>(93.26%, 94.27%) | 95.63%<br>(95.19%, 96.05%) |
| EHR-Derived + Third-Party Mortality + Third-Party Claims | 81.79%<br>(81.05%, 82.52%) | 94.82%<br>(94.26%, 95.33%) | 96.19%<br>(95.78%, 96.57%) | 76.51%<br>(75.59%, 77.42%) | 86.11%<br>(85.37%, 86.82%) | 93.78%<br>(93.26%, 94.27%) | 95.61%<br>(95.17%, 96.02%) |

### Subgroup Analyses

Sensitivity was stable for the following stratification variables, with all subgroups having sensitivity of 80% or greater: year of diagnosis (80.20-83.36%), number of recorded lines of therapy (80.91-83.02%), and sex (female: 80.92%, male: 82.77%). Age at diagnosis showed greater variation, with younger patients having lower sensitivity than older patients (e.g., <35: 73.60%, ≥75: 86.32%). Cancer type differences were also observed, with BC (78.41%) and CRC (79.04%) as the lowest and NSCLC (84.03%), OC (80.66%), PANC (83.84%), and PC (81.93%) all above 80%. Geographic differences were also apparent, with the West (69.04%) lower than other regions (Midwest: 86.48%, Northeast: 78.17%, South: 83.63%, Other/Missing: 78.76%). Sensitivity also differed by race (Asian: 70.07%, Black or African American: 77.90%, White: 84.79%, Other Race: 81.01%, Missing: 78.77%). Specificity was at or above 94% for all groups except age ≥75 (91.56%), NSCLC (93.68%), PANC (84.81%), and South (93.77%). Exact date agreement was stable across all stratification variables (84.16-89.62%). Date agreement metrics were higher for all stratification variables with a 15 day tolerance (91.21-94.98%) and a 30 day tolerance (94.31-97.32%).

We also evaluated sensitivity and date agreement metrics stratified by the year of death in NDI. Sensitivity was lowest for deaths in 2021 (80.61%) and 2022 (78.26%). Exact date agreement was stable by NDI year of death (88.80-90.16%), as well as with a 15 day tolerance (97.14-98.05%) and a 30 day tolerance (97.30-99.75%). Full subgroup analysis results are shown in **Table 3** (please see end of document).

**Table 3:**
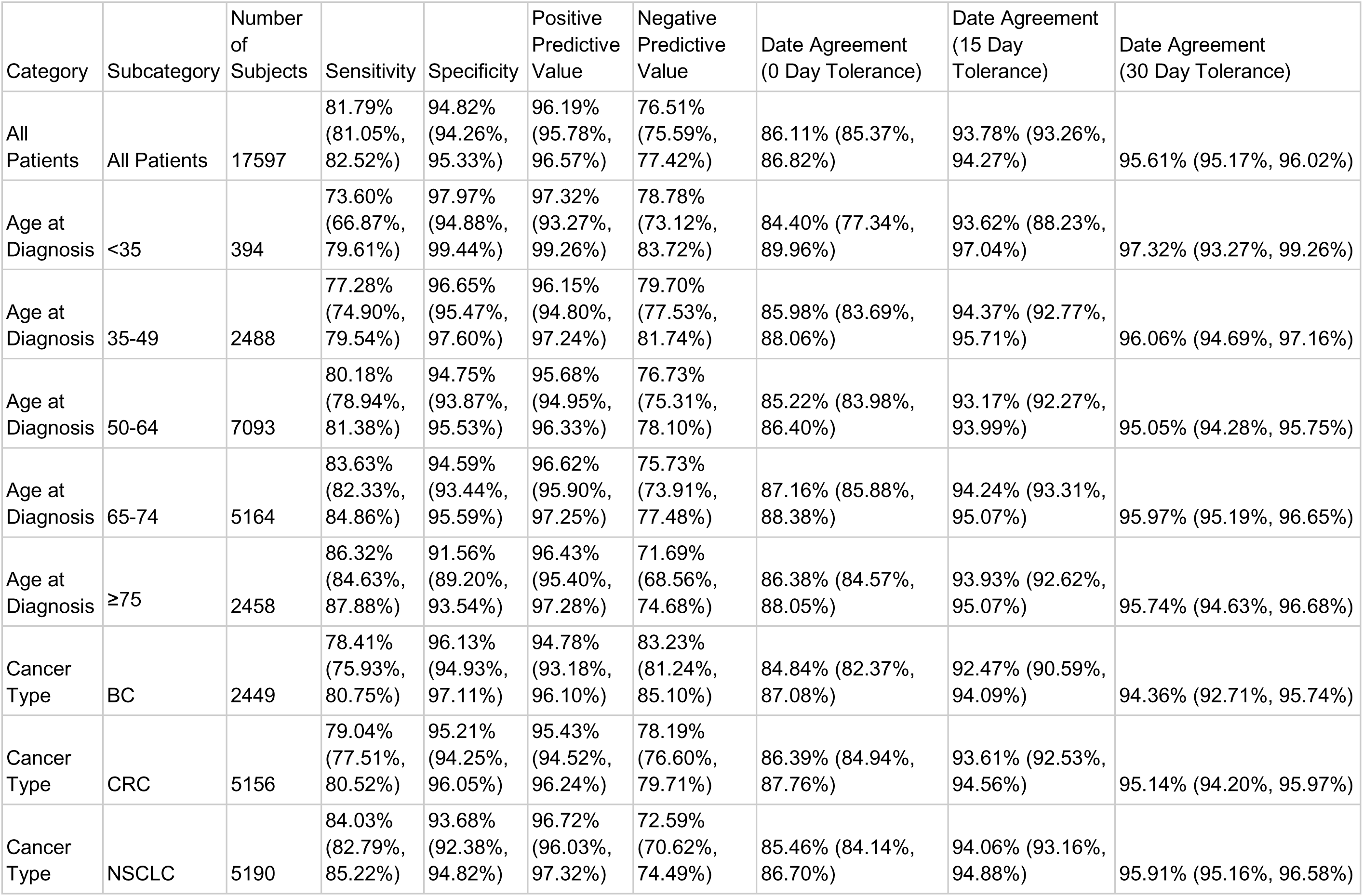
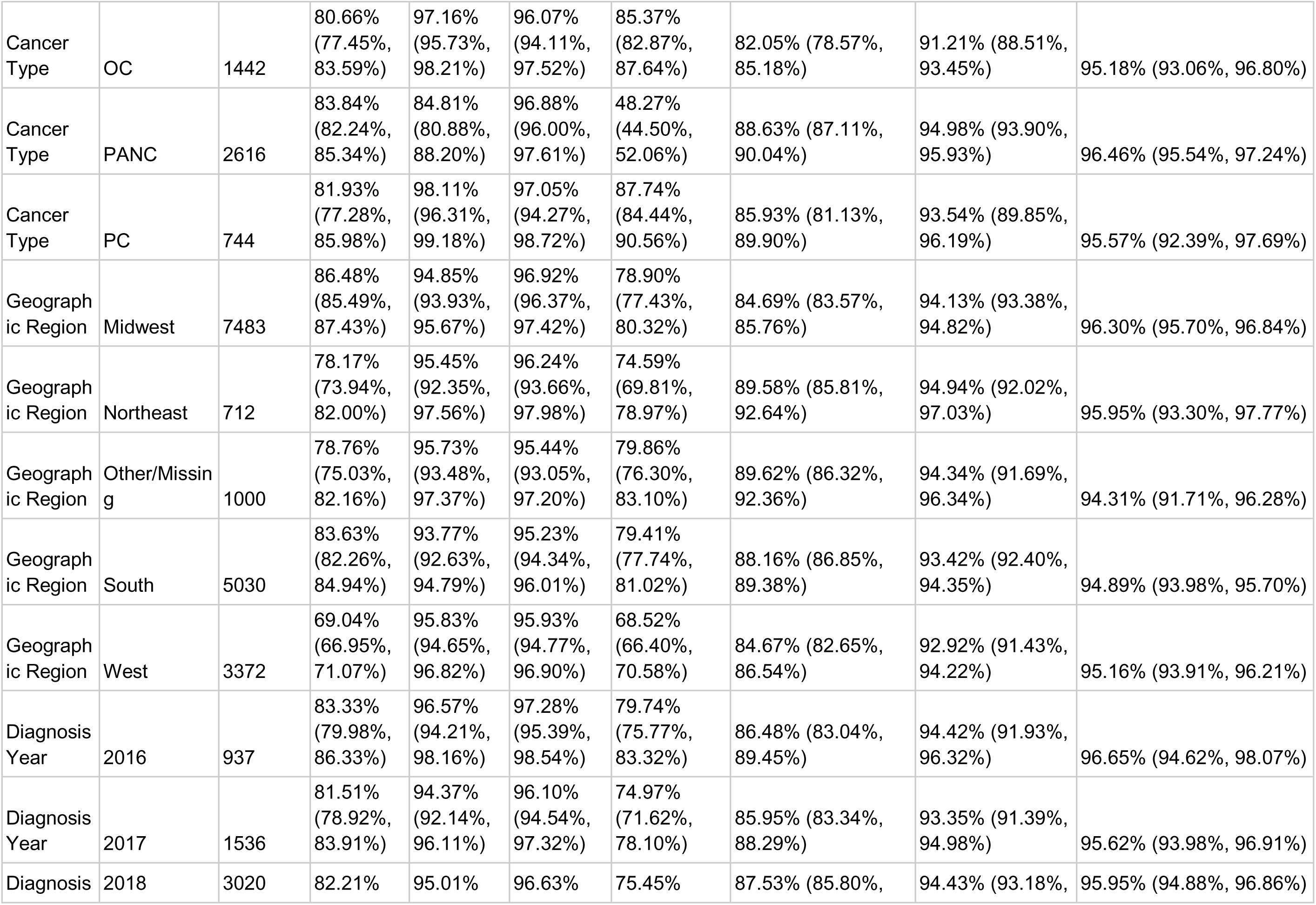
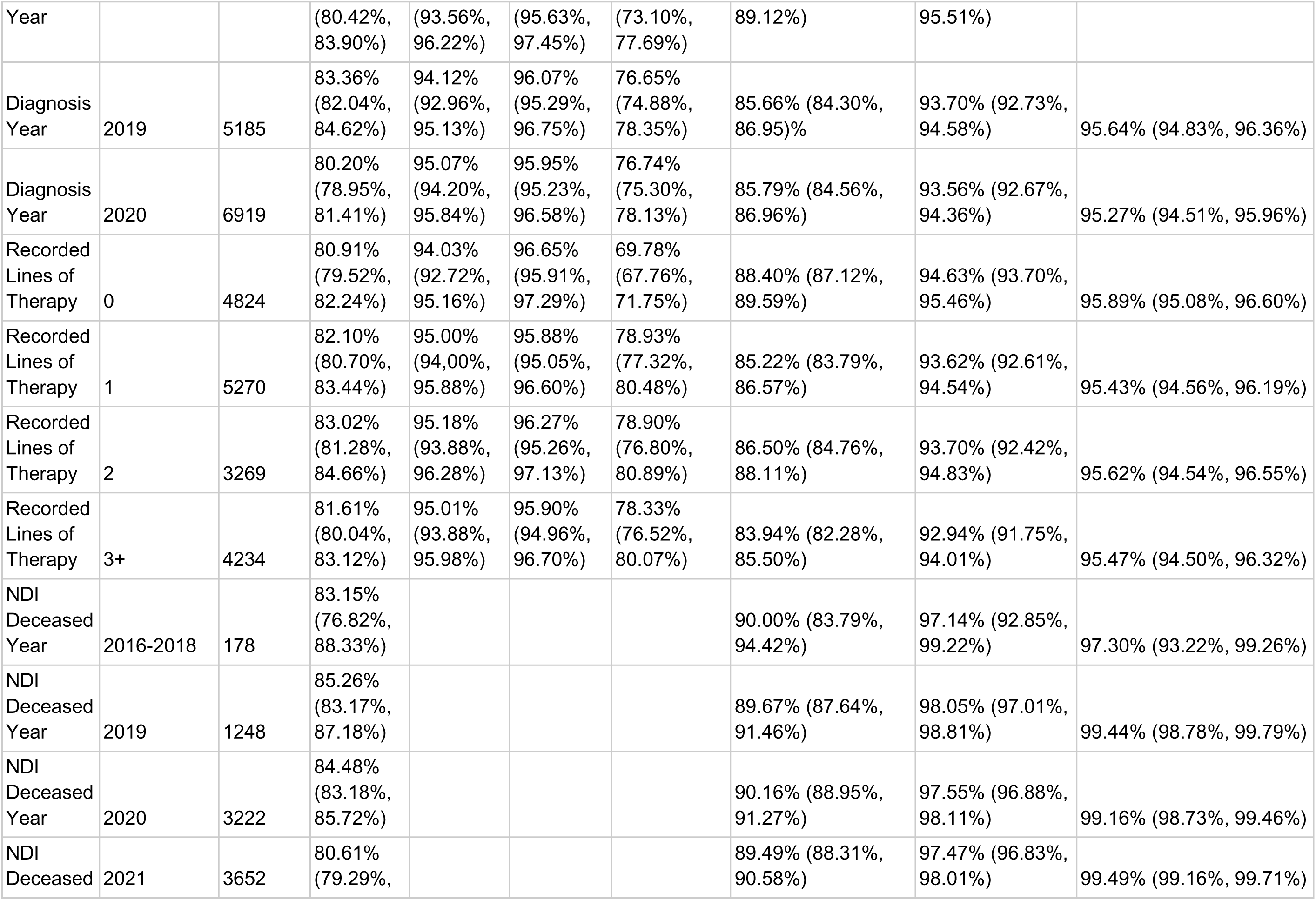
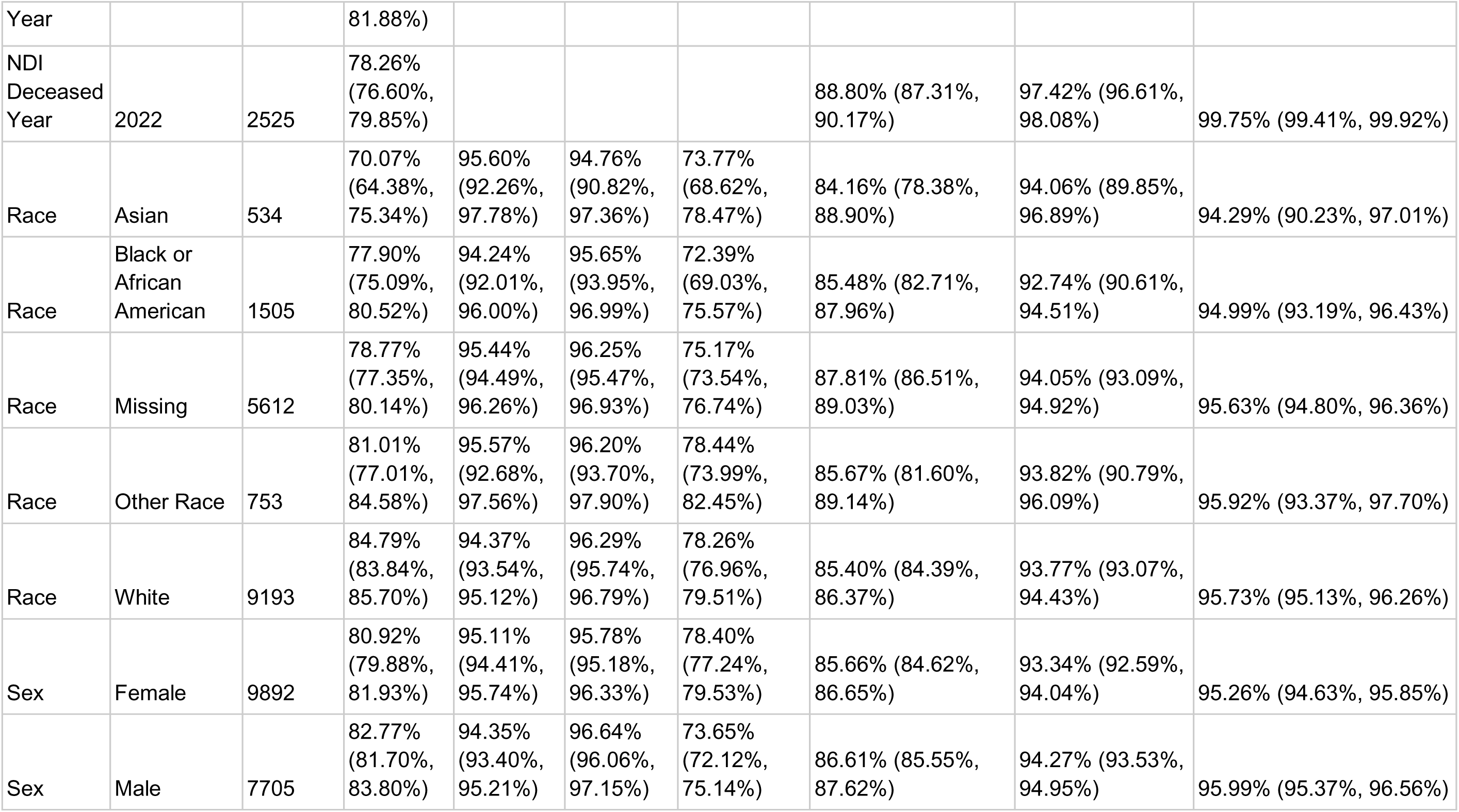
Performance Metrics by Subgroup

### Cumulative Cases / Dynamic Controls (Table 4)

When excluding patients lost to follow-up at time t, sensitivity was 95.95% at t=6 months, 96.57% at t=12 months, and 97.96% at t=24 months. Specificity was 99.64% at 6 months, 99.29% at 12 months, and 98.25% at 24 months.

**Table 4:** Cumulative Cases and Dynamic Controls

| Timeframe | Sensitivity | Specificity |
| --- | --- | --- |
| 6 Months | 95.96% (94.96%, 96.82%) | 99.64% (99.53%, 99.73%) |
| 12 Months | 96.57% (95.88%, 97.16%) | 99.29% (99.13%, 99.43%) |
| 24 Months | 97.96% (97.56%, 98.31%) | 98.25% (97.96%, 98.50%) |

## Discussion

Real-world databases are valuable research tools for precision oncology. The ability to capture rich, longitudinal clinical data within these databases is crucial for investigating the relationships between molecular features, treatment responses, and patient outcomes. The reliability of overall survival analyses conducted using such data is dependent on the quality of the mortality data captured.

In this study, we assessed the performance of the Tempus composite mortality variable by benchmarking it against the NDI. Our findings indicate that the composite mortality variable captures 82% of death events recorded in the NDI. We observed increased sensitivity when using the cumulative cases and dynamic controls approach, suggesting that subjects with deaths not captured by the Tempus variable are censored appropriately rather than misclassified as alive. The high positive predictive value (PPV) observed indicates that when the composite mortality variable identifies a patient as deceased, this classification is highly reliable. Date agreement analyses showed strong concordance with NDI-reported dates of death, with an exact agreement of 86%. This agreement increased when allowing for a ±15-day and ±30-day tolerance window. Collectively, these results suggest that while the Tempus composite mortality variable may not capture every death event, the death events it does report are highly dependable, and patients with uncaptured death events are correctly categorized as lost to follow-up rather than misclassified as alive.

Consistent with previous research (5,12), we observed that the addition of external mortality data sources dramatically improved sensitivity to levels suitable for outcomes research. A substantial portion of this sensitivity improvement is attributed to the inclusion of third-party mortality data from Veritas, which itself aggregates information from various mortality sources, including government records, obituary notices, and public cemetery records. We also observed a modest additional increase in sensitivity with the inclusion of third-party claims data, although mortality data are not the primary rationale for integrating claims into the Tempus database.

Much of the variability observed across subgroups in our study aligns with patterns reported in prior investigations. For example, results from Zhang et al. (12) similarly show BC and CRC among the cancer types with the lowest sensitivities, NSCLC and PANC among the highest, and sensitivity increasing with patient age. Given these similarities, we hypothesize that this variability may be driven by differences in expected survival. Patients with longer survival require extended follow-up, which increases their chance of being lost to follow-up from the source EHR. Subsequently, locating these patients in supplementary external datasets becomes more challenging over time, as the extended period increases the likelihood of personal data changes that complicate record linkage. Therefore, reduced sensitivity in these groups may stem from this dual challenge of patient attrition from the EHR and a diminished ability to match identifiers across external sources.

Expected survival also appears to influence specificity, a hypothesis supported by two patterns observed in both our study and in Zhang et al (12): PANC had the lowest specificity, and specificity consistently declined with increasing patient age. While the NDI is the gold standard, it may not capture every death. In a cohort with a very high mortality rate like PANC, any true death missed by the NDI but captured by a real-world data source would be misclassified as a false positive, thereby artificially lowering the reported specificity. Though not all false positives in our study are NDI errors, this effect could impose an upper limit on achievable specificity in shorter-lived populations.

While the expected survival hypothesis may explain many observed patterns, other factors likely contribute to the variability in specific subgroups. For instance, consistent with previous research (4,12), sensitivity was lower among patients from several racial and ethnic minority groups, including Asian and Black or African American patients, compared to White patients.

For some populations, there may be challenges in harmonizing patient names across different cultural and linguistic contexts, including variations in name structure and the use of multiple given names, which complicate patient identification and record linkage across datasets.

Similarly, our observation of lower sensitivity in the Western U.S. may be influenced by other factors. Regional differences in end-of-life practices could contribute to this disparity. For instance, variations in where death occurs (e.g., in-hospital versus at home), customs for death notification, or the prevalence of cremation over burial could impact data capture. Ultimately, these specific variations highlight the complex interplay of demographic and cultural factors and underscore the need for continued refinement of methods for real-world data collection and data linkage.

This study has several limitations. First, the primary objective was to validate the mortality endpoint specifically for use within the Tempus multimodal database. Therefore, the study population was intentionally limited to patients who received Tempus NGS testing. While this design is essential for the intended use-case of enabling molecularly driven outcomes analysis, it means the cohort may not be fully representative of the general oncology population, which could limit the generalizability of our specific performance findings. Second, while we have proposed hypotheses for the observed variations in sensitivity and specificity, this study was not designed to definitively determine their causal factors. Third, our analysis was based on data with a cut-off date of 2022. More recent data, which would reflect increases in Tempus’s sequencing volume over time and evolving data capture methods, could potentially yield different performance metrics. Future studies utilizing more contemporaneous cohorts would be valuable to assess the evolving performance of the Tempus composite mortality variable. Despite these limitations, this study successfully validates the Tempus composite mortality variable against the NDI gold standard for use in real-world outcomes research.

## Conclusions

This study validates the Tempus composite mortality variable as a robust and reliable endpoint for conducting real-world evidence analyses. The high accuracy of identified deaths and the appropriate censoring of patients lost to follow-up support its use in overall survival analyses. Validating the quality of this mortality data is a foundational step for the broader goal of the Tempus multimodal dataset: enabling high-quality research to improve patient outcomes and advance cancer drug development.

## Author Contributions

JK: Conception, study design, data acquisition, data analysis, manuscript drafting

FI: Study design

ER: Data acquisition

JD: Conception, study design

SM: Data acquisition

ES: Manuscript drafting

SWH: Study design

CS: Conception, study design, manuscript drafting

All authors have reviewed and approved the final version of this manuscript.

## Acknowledgments

The authors thank Jesus Roman for building data pipelines and managing external data submissions and retrievals; Dongho Shin for verifying our analytical code; and Shannon Mulmat for programming support. We thank Shahir Kassam-Adams and Kirsty Macaulay for their support in integrating Veritas mortality data and reviewing study results. We thank Dana DeSantis from the Tempus Scientific Communications Team for writing support.

## Declarations

All authors are current or former employees of Tempus AI, Inc. and may hold stock in the company. There is no external funding associated with this work.

## List of Abbreviations

CC/DC: cases/dynamic controls
EHRs: electronic health records
CDC: Centers for Disease Control
NDI: National Death Index
SSA LADMF: Social Security Administration’s Limited Access Death Master File
NGS: Next-generation sequencing
NPV: negative predictive value
PII: personally identifiable information
PPV: positive predictive value
SSDI: Social Security Death Index
RWD: real-world data
RWE: real-world evidence
NSCLC: non-small cell lung cancer
BC: breast cancer
OC: ovarian cancer
PANC: pancreatic cancer
CRC: colorectal cancer
PC: prostate cancer

